# Bayesian Spatio-Temporal Modelling of Reported Terminated Pregnancy Across Nigerian States (2013-2024)

**DOI:** 10.64898/2026.02.16.26346435

**Authors:** Taiwo Oyewale Asifat, Oluwafemi Lawal Bisiriyu, Abolaji Moses Ogunetimoju

## Abstract

**Introduction:** The long-standing disconnection of abortion legislation in Nigeria with the estimated incidence of 1.8 million terminations a year has contributed to systematic gaps in reliable abortion data for health policy. Any subnational monitoring under conditions of legal restraint tends to remain hidden beneath under-reporting and spatial instability such that policy makers are not left with a clear picture of where and why these decisions are being made.

**Methods and Analysis:** To address this ambiguity, this paper traces the path of state-level evolution of reproductive choices within the 2013, 2018 and 2024 NDHS. We detected the latent socio-demographic causes of terminated pregnancy using a Bayesian spatio-temporal framework, such as wealth, education, literacy, and contraceptive prevalence.

**Results:** The rates were highly spatio-temporally intense and polarized in the region, with probabilistic evidence to justify state-specific reproductive health interventions between 2013 and 2024. Southern and coastal states (e.g., Lagos, Bayelsa) demonstrated sustained increases in prevalence in line with a high fertility transition, termination is more reproductive agency, access to services and reporting. Conversely, the unmet contraceptive need and structural vulnerability were the major causes of increased rates in the northern states (e.g., Yobe, Kano). Patterns of determinants also changed with time: in previous surveys, household wealth turned out to be a protective factor, as of 2024, education and literacy had become the strongest predictors.

**Conclusions:** Such findings affirm a dual reproductive regime in Nigeria—choice based in the South and vulnerability based in the North necessitating a shift from homogenous national approaches to state-specific reproductive health policies.

*What is already known on this topic:* Studies have noted the continuous disparities in the maternal and reproductive health indicators between northern and southern Nigerian states. Nevertheless, most of the studies done before were based on cross-sectional analysis and national-level summaries. Not many considered spatial dependence among states or studied how decisions on termination vary over time.

*What this study adds:* Through shared modelling of spatial effects, temporal trends and space-time interactions, it establishes consistent high-risk conditions, arising hotspots and areas with decoding risk. The Bayesian model enhances the accuracy of the estimation since it takes into consideration the spatial correlation and the strength of borrowing on neighboring states.

*How this study might affect research, practice or policy:* In the case of research, the study offers a methodological approach to the analysis of other maternal and public healthcare indicators by small-area estimation methods. Practically, with high-risk and emerging hotspots states identified, reproductive health more focused interventions can be implemented and limited resources can be efficiently allocated. To the policy, the study provides state-specifics evidence to inform subnational reproductive health planning and monitoring.

## 1.0 INTRODUCTION

Unsafe abortion is a top cause of maternal deaths in most parts of the world, burdened more by Sub-Saharan Africa. This is an urgent social health issue because restrictive legal frameworks and inadequate access to the full range of sexual and reproductive health services contribute to high morbidity and mortality rates (Zegeye et al., 2024). Nigeria, the most populous country in the region, is the embodiment of this crisis. Approximately 1.8 million induced abortions are performed each year under a legal code that only authorizes the practice to protect the life of the mother (Federal Republic of Nigeria, 1990; Hussain et al., 2018). It associated with severe complications and an estimated mortality rate of about 212 deaths per 100,000 cases due to this contradiction between the prevalence of practice and harsh restriction, which ends terminations in secret locations (Akinlusi et al., 2018). Their drivers are multidimensional: whereas 40 percent of pregnancies all over the world are untimed, 61 percent of them are terminated by abortion (Bearak et al., 2020), in Nigeria, the situation is aggravated by the fact that modern contraceptive prevalence rate (MCPR) stagnates at 10-12 percent (Abubakar et al., 2024), especially in the north, where contraceptive use is limited by sociocultural and religious beliefs.

Nigeria reproductive health landscape is highly heterogenous but are characterized by a deep spatial heterogeneity. Well-established sociocultural customs, gross economic inequalities, and a very uneven healthcare system make the issue of risk a mosaic of complication in its 36 states and the Federal Capital Territory (Ibeji et al., 2022). Research findings support the presence of strong regional disparities, where North East and North Central areas have high rates of abortion due to acute lack of services, but southern areas, despite their relative infrastructural improvements, still have a significant number of challenges (Okonofua et al., 2021). The Nigeria Demographic and Health Surveys (NDHS) are very important in this matter, but there is a severe gap in methodology. Conventional analytic methods, such as standard regression models used in research, such as Oye-Adeniran et al. (2020), often neglect two basic data structures: spatial dependency, where the outcomes of any one geographic unit are inherently connected to the outcomes of the neighboring units, and temporal evolution, which is critical to the formation of the trends in response to changing socioeconomic and policy environments (Tolani et al., 2024). This means that predetermined estimates are usually weak, obscured, and not stable in data-sparse regimes, which restricts their application to specific intervention.

The developments made in spatial epidemiology have recently started tackling these complexities. The Ibeji et al. (2022) and Nnanatu et al. (2022) studies have indeed proven that Bayesian hierarchical models are useful to estimate small areas of health indicators in Nigeria by demonstrating how these models can take advantage of space and time by “borrowing strength” to stabilize the estimate. Also, as noted in emerging research by Oluwafemi et al. (2025), unsupervised classification can be useful in disclosing latent spatial patterns in health crises. Nevertheless, no synthesis and concrete use of these sophisticated methodologies on the delicate, underreported area of terminated pregnancy choices have been provided. Current literature has not successfully incorporated a spatio-temporal process, which models risk continuously throughout the Nigerian domain, and quantifies uncertainty rigorously-which is of paramount significance as abortion is a stigmatized and clandestine activity. This gap is directly taken care of in this study. We are operationally inspired by the approach used by Ibeji et al. (2022) as well as the covariance frameworks presented by Ying-qing et al. (2023), which are applied to construct a customized Bayesian spatio-temporal model. Our model goes beyond clustering to give a probabilistic rigorous, continuous surface of risk estimation, which is formally structured to address underreporting and sparsity of spatial data as observed by Opeyemi et al. (2023).Thus, the study objectives include: (1) to simulate the spatio-temporal distributions of terminated pregnancy choices by state in 2013, 2018 and 2024, identifying persistent and emergent high-risk groups; (2) to quantify the scale and importance of state-level changes and (3) to evaluate the relative controller of the identified spatial disparity, the key socioeconomic, demographic, and access to healthcare factors. This study contributes to the methodological improvement by explicitly treating the problem of spatial dependence and data sparseness present in the previous analyses, as it applies an integrated Bayesian hierarchical model with a Gaussian random field and Matern covariance function (Ibeji et al., 2022; Opeyemi et al., 2023). The results have a high level of applied importance, as they provide accurate, evidence-based hotspots maps that define how the Federal Ministry of Health (2021) and partner non-governmental organizations should allocate resources and choose more specific programmatic approaches, which could contribute to the achievement of Sustainable Development Goal (SDG) 3.7 directly. Though we are cognizant of the inherent limitations of data (such as underreporting bias and the spatial displacement of NDHS cluster coordinates), our methodology uses Bayesian small area estimation as the most robust analytical model currently in existence that can be used to generate plausible subnational indicators on such a sensitive subject. Finally, this paper provides a replicable framework of geospatial analysis of reproductive health outcomes in other oppressive legal settings in West Africa.

## 2.0 METHODOLOGY

This research uses a secondary source of information that is based on Nigeria Demographic and Health Surveys (NDHS) which were carried out in 2013, 2018 and 2024 respectively. The NDHS is a nationally representative, cross-sectional household survey, which is conducted by the National Population Commission in partnership with the DHS Program, and it is the most detailed source of reproductive health information in Nigeria. The survey uses a multi-stage cluster stratified sampling design to provide representativeness in terms of states and geopolitical zones and urban-rural environments. To make this analysis, the individual-level reproductive histories were pulled out among the women aged 15-49 years, paying close attention to the self-reported events concerning the unplanned pregnancy and terminated pregnancy choices. These data points were commonly adopted as they are the best proxies of abortion-related behavior under the DHS framework due to legal constraints on abortion reporting in Nigeria and its sensitivity. The explanatory variables were selected based on known theory of reproductive health and previous empirical findings and contained socioeconomic variables (education, literacy and household wealth index) and health service use variables (contraceptive use and utilization of antenatal care).

In order to conduct spatial analysis, the NDHS geospatial datasets were used to get the geographic coordinates of survey clusters. According to DHS confidentiality, in urban regions, GPS coordinates are shifted randomly within 2 km, whereas in rural regions, it is shifted within 5 km with a small percentage shifting up to 10 km. Although this introduce spatial uncertainty at fine scale, it is still suitable to state level analysis. Spatial adjacency structures were defined by the Administrative Boundary shapefiles of 36 states and the Federal Capital Territory of Nigeria, derived out of the Global Administrative Areas (GADM) database.

### 2.1. Statistical Modelling Strategy

The analytical purpose of this work was to measure both spatial and temporal fluctuation in rates of terminated pregnancies in Nigeria whilst taking into consideration unobserved spatial dependence, time dynamics, and socioeconomic heterogeneity. Our model is based on a Bayesian hierarchical model of modelling, which is ideally applicable in reproductive health outcomes, whose characteristics are underreporting, spatial clustering, and data density. The proportion of pregnancies terminated each state-year by DHS was recalculated into counts of the terminated pregnancies by multiplying the prevalence estimates by the number of surveyed women per state-year, providing the ability to model counts but preserving the structure of the survey.

Through the aggregation of the data and the emphasis on relative risk estimation, we used a Poisson-based Besag-York-Mollie (BYM) model instead of a binomial formulation. The Poisson-BYM model permits observed values to be modeled as a proportion of the expected values, supports heterogeneity in population across states, and explicitly represents both population spatial autocorrelation and unstructured variability. The fact that it can be extended to spatio-temporal allows it to be specifically used to analyze alterations in the 2013 survey wave, the 2018 survey wave, and the 2024 survey wave. We have developed a modelling approach that has gone through in three layers.

First, the overall changes in terminated pregnancy risk by survey years were studied through temporal decomposition. Second, spatial random effects were considered to determine the clustering and smooth estimates of risks among neighboring states at the state level. Third, the complete spatio-temporal hierarchical model was allowed to fit simultaneously relative risks, controlling the spatial proximity and temporal changes. The socioeconomic and health service variables were included as fixed effects to determine their relationship with risk of terminated pregnancy after consideration of latent spatial and temporal structure.

### 2.2. Mathematical Specification of the Poisson–BYM Model

Let *y_it_*denote the observed number of women reporting a terminated pregnancy in state during year. When individual counts were unavailable, these were reconstructed as

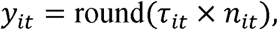

where *τ_it_*is the state–year terminated pregnancy rate and *n_it_*is the number of surveyed women. Expected counts were defined as

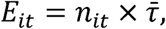

Where,

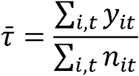

represents the national average terminated pregnancy rate across all states and years.

We assumed a Poisson likelihood with a log-offset:

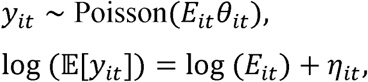

where the linear predictor is given by

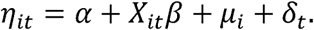

Here, α is the intercept, *X_it_* is a vector of standardized covariates, and *β* represents their corresponding regression coefficients. The spatial effect μ_i_ captures unobserved state-level heterogeneity, while δ_t_ models The spatial effect was decomposed using the classical BYM formulation:

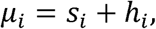

where *s*_i_is a structured spatial component modeled using an intrinsic conditional autoregressive (ICAR) prior, and *h*i is an unstructured random effect capturing state-specific noise.

The ICAR component was specified as:

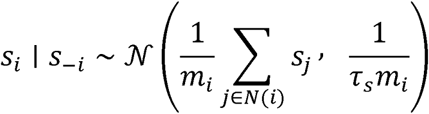

where N (*i*)denotes the set of neighboring states of state i,m_i_ is the number of neighbors, and τ_s_is the precision parameter.

We used weakly informative priors throughout and followed common INLA recommendations:

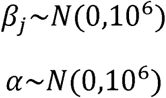

and Gamma or PC priors for precision parameters (default INLA *Gamma*(1,10)^5^ on precision or PC priors for BYM2 mixing and total precision). The structured ICAR component satisfies the conditional prior

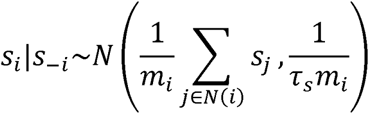

Where:

m_i_=|N(i)|= the number of neighbours of area i

*N(i)*= set of neighbors of i

*s_i_* spatial random effect for area i

*τ_s_*precision _inverse variance parameter

**Note: This is the intrinsic conditional autoregressive (ICAR) prior underlies the BYM/BYM2 model.**

Posterior summaries for β (log-relative-risk scale) were exponentiated to yield relative risks (RRs).

### Ethical Consideration

This study is a secondary analysis of de-identified datasets from the 2013, 2018, and 2024 Nigeria Demographic and Health Surveys (NDHS). Ethical approval for the original surveys was obtained by the National Population Commission (NPC) from the National Health Research Ethics Committee of Nigeria (NHREC) and the Institutional Review Board of ICF International. The authors obtained permission from the DHS Program to use the data. This research was conducted in accordance with the Declaration of Helsinki; because it utilizes completely anonymized, publicly available secondary data, it was exempt from further institutional ethical review.

## 3.0 RESULTS

Table 1 shows that patterns of terminated pregnancies differ significantly in space and time across Nigerian states between 2013 and 2024, which are highly correlated with gradients in contraceptive use, educational attainment and household wealth. It is worth noting that increased rates of higher terminated pregnancy were consistently recorded in socioeconomically advantaged states and mostly southern states such as Lagos, Imo, Anambra, Akwa Ibom, Ekiti, Osun, Ogun, Delta, Rivers, and Bayelsa which continued to rise by 2024. This trend questions the deprivation-based explanations of abortion and offers, instead, an explanation of its use as an adjunctive fertility regulation measure in situations where women are more educated, have later onset of childbearing, and have increased reproductive choice. Lagos offers a good example here: the terminated pregnancy percentages were steady and consistent within all waves of the survey regardless of contraceptive prevalence being more than 50 percent which suggests that abortion is complementary to, but not substitutive of, modern contraception. Likewise, the dramatic changes in states like Ekiti, Osun, Bayelsa, Imo, and Akwa Ibom by 2024 were also accompanied by the improvement in education and uptake of contraceptives, supporting the view that the terminated pregnancy is a deliberate fertility management strategy and not a sign of the socioeconomic disadvantage. Together, these results point to the significance of putting the terminated pregnancy into the context of more general reproductive patterns organized by socioeconomic transition, and not seeing it as simply being a consequence of disadvantage. Northern states, on the contrary, such as Kano, Kaduna, Bauchi and Sokoto, had an increasing or unstable termination rates with continuing low contraceptive coverage, education and wealth. This deviation indicates elevated degrees of unmet requirement and constrained reproductive control since termination rates frequently shot up even wherever contraceptive utilization merely enhanced. There were rapid growths in the areas of termination of pregnancies in conflict prone areas such as Borno, Yobe and Adamawa with lack of corresponding improvements in the socioeconomic parameters thus suggesting that there is a crisis-based trend. On the other hand, an isolated team comprising of Kwara, Plateau and Rivers reported falling or stable figures by 2024, probably because of increased fertility management or regionalized changes in reproduction.

**Table 1:** Socio-Demographic Characteristics of the Study Respondents.

| State | Year | N | Abortion rate (%) | Contraceptive use rate (%) | Avg education | Avg wealth |
| --- | --- | --- | --- | --- | --- | --- |
| Abia | 2013 | 469 | 8.96 | 36.7 | 2.95 | 3.97 |
| Abia | 2018 | 358 | 4.19 | 13.7 | 3.04 | 4.30 |
| Abia | 2024 | 470 | 15.5 | 35.7 | 3.16 | 4.16 |
| Adamawa | 2013 | 901 | 11.1 | 4.11 | 1.87 | 2.66 |
| Adamawa | 2018 | 548 | 8.21 | 30.3 | 1.69 | 2.45 |
| Adamawa | 2024 | 789 | 25.9 | 19.4 | 1.89 | 2.05 |
| Akwa Ibom | 2013 | 533 | 5.25 | 24.0 | 2.68 | 3.49 |
| Akwa Ibom | 2018 | 307 | 13.7 | 20.5 | 2.86 | 3.27 |
| Akwa Ibom | 2024 | 494 | 28.3 | 40.9 | 2.95 | 3.61 |
| Anambra | 2013 | 491 | 11.8 | 35.6 | 2.84 | 4.09 |
| Anambra | 2018 | 504 | 5.56 | 48.4 | 2.93 | 4.04 |
| Anambra | 2024 | 643 | 26.3 | 40.1 | 3.07 | 4.32 |
| Bauchi | 2013 | 1259 | 18.0 | 2.94 | 1.36 | 1.91 |
| Bauchi | 2018 | 799 | 16.6 | 8.14 | 1.51 | 1.98 |
| Bauchi | 2024 | 884 | 12.2 | 10.4 | 1.44 | 1.99 |
| Bayelsa | 2013 | 763 | 11.0 | 13.6 | 2.51 | 3.80 |
| Bayelsa | 2018 | 337 | 5.93 | 5.34 | 2.67 | 3.47 |
| Bayelsa | 2024 | 513 | 31.2 | 23.8 | 2.89 | 3.69 |
| Benue | 2013 | 599 | 14.5 | 17.7 | 2.29 | 2.59 |
| Benue | 2018 | 523 | 7.84 | 13.0 | 2.40 | 2.57 |
| Benue | 2024 | 631 | 18.7 | 29.6 | 2.51 | 2.68 |
| Borno | 2013 | 572 | 6.99 | 1.75 | 1.46 | 2.59 |
| Borno | 2018 | 587 | 9.03 | 2.90 | 1.49 | 2.49 |
| Borno | 2024 | 671 | 10.7 | 9.69 | 1.65 | 2.12 |
| Cross River | 2013 | 490 | 12.4 | 31.0 | 2.53 | 3.20 |
| Cross River | 2018 | 248 | 16.5 | 21.0 | 2.78 | 3.10 |
| Cross River | 2024 | 549 | 21.3 | 29.0 | 2.89 | 3.36 |
| Delta | 2013 | 646 | 5.12 | 32.8 | 2.53 | 3.87 |
| Delta | 2018 | 292 | 6.16 | 14.7 | 2.82 | 3.90 |
| Delta | 2024 | 613 | 16.0 | 37.2 | 3.03 | 4.10 |
| Ebonyi | 2013 | 628 | 17.4 | 18.5 | 2.31 | 2.60 |
| Ebonyi | 2018 | 565 | 10.1 | 9.56 | 2.55 | 2.69 |
| Ebonyi | 2024 | 840 | 18.7 | 10.6 | 2.60 | 2.66 |
| Edo | 2013 | 568 | 8.45 | 31.0 | 2.67 | 3.99 |
| Edo | 2018 | 249 | 11.6 | 16.9 | 2.80 | 3.59 |
| Edo | 2024 | 541 | 8.87 | 27.9 | 3.00 | 4.44 |
| Ekiti | 2013 | 497 | 11.5 | 27.0 | 3.13 | 4.27 |
| Ekiti | 2018 | 279 | 12.9 | 38.4 | 2.95 | 3.57 |
| Ekiti | 2024 | 369 | 26.0 | 62.9 | 3.14 | 3.93 |
| Enugu | 2013 | 528 | 11.7 | 37.3 | 2.74 | 3.56 |
| Enugu | 2018 | 332 | 6.93 | 38.6 | 2.97 | 3.50 |
| Enugu | 2024 | 485 | 14.2 | 25.4 | 2.82 | 3.84 |
| Gombe | 2013 | 1029 | 14.6 | 4.96 | 1.50 | 2.08 |
| Gombe | 2018 | 701 | 19.8 | 22.4 | 1.56 | 2.12 |
| Gombe | 2024 | 806 | 18.6 | 32.0 | 1.78 | 2.20 |
| Imo | 2013 | 437 | 18.3 | 38.9 | 3.01 | 3.87 |
| Imo | 2018 | 378 | 18.0 | 35.7 | 3.04 | 3.84 |
| Imo | 2024 | 653 | 27.3 | 45.3 | 3.16 | 4.05 |
| Jigawa | 2013 | 1242 | 10.5 | 0.64 | 1.18 | 1.72 |
| Jigawa | 2018 | 754 | 19.5 | 5.17 | 1.34 | 1.79 |
| Jigawa | 2024 | 955 | 12.4 | 4.92 | 1.37 | 1.62 |
| Kaduna | 2013 | 826 | 10.3 | 20.0 | 1.81 | 2.94 |
| Kaduna | 2018 | 738 | 16.0 | 17.2 | 1.83 | 2.92 |
| Kaduna | 2024 | 1080 | 22.2 | 13.9 | 2.06 | 2.78 |
| Kano | 2013 | 1937 | 7.23 | 0.72 | 1.48 | 2.43 |
| Kano | 2018 | 1054 | 13.0 | 6.26 | 1.63 | 2.50 |
| Kano | 2024 | 1210 | 23.6 | 12.1 | 1.89 | 2.88 |
| Katsina | 2013 | 1321 | 7.38 | 1.82 | 1.31 | 2.09 |
| Katsina | 2018 | 832 | 15.9 | 4.33 | 1.55 | 2.42 |
| Katsina | 2024 | 914 | 12.1 | 8.86 | 1.70 | 2.53 |
| Kebbi | 2013 | 1043 | 7.00 | 1.53 | 1.15 | 1.84 |
| Kebbi | 2018 | 660 | 11.8 | 4.24 | 1.20 | 2.03 |
| Kebbi | 2024 | 1008 | 19.7 | 4.37 | 1.23 | 1.62 |
| Kogi | 2013 | 462 | 4.33 | 11.3 | 2.51 | 3.57 |
| Kogi | 2018 | 320 | 13.8 | 18.4 | 2.64 | 3.21 |
| Kogi | 2024 | 699 | 8.30 | 13.2 | 2.36 | 2.81 |
| Kwara | 2013 | 637 | 3.92 | 38.0 | 2.19 | 3.89 |
| Kwara | 2018 | 380 | 4.47 | 19.5 | 2.26 | 3.01 |
| Kwara | 2024 | 664 | 1.51 | 11.1 | 2.12 | 3.07 |
| Lagos | 2013 | 885 | 17.9 | 53.8 | 2.99 | 4.81 |
| Lagos | 2018 | 445 | 17.8 | 53.9 | 3.11 | 4.75 |
| Lagos | 2024 | 631 | 18.2 | 56.9 | 3.28 | 4.83 |
| Nasarawa | 2013 | 597 | 9.73 | 19.3 | 2.03 | 3.01 |
| Nasarawa | 2018 | 453 | 8.61 | 14.8 | 2.31 | 3.24 |
| Nasarawa | 2024 | 750 | 31.2 | 26.8 | 2.24 | 3.44 |
| Niger | 2013 | 899 | 3.68 | 7.34 | 1.51 | 2.84 |
| Niger | 2018 | 666 | 15.5 | 8.71 | 1.61 | 2.72 |
| Niger | 2024 | 935 | 6.20 | 13.0 | 1.59 | 2.77 |
| Ogun | 2013 | 499 | 13.4 | 27.1 | 2.34 | 3.93 |
| Ogun | 2018 | 293 | 6.48 | 33.8 | 2.53 | 3.95 |
| Ogun | 2024 | 670 | 22.1 | 41.8 | 2.84 | 4.24 |
| Ondo | 2013 | 600 | 13.5 | 27.7 | 2.54 | 3.53 |
| Ondo | 2018 | 299 | 4.35 | 18.4 | 2.77 | 3.29 |
| Ondo | 2024 | 483 | 11.6 | 23.6 | 2.77 | 4.15 |
| Osun | 2013 | 537 | 3.72 | 35.6 | 2.81 | 4.14 |
| Osun | 2018 | 278 | 11.5 | 24.8 | 2.83 | 3.54 |
| Osun | 2024 | 434 | 28.8 | 41.5 | 3.11 | 4.26 |
| Oyo | 2013 | 625 | 11.7 | 34.4 | 2.28 | 3.50 |
| Oyo | 2018 | 367 | 6.54 | 19.6 | 2.64 | 3.76 |
| Oyo | 2024 | 745 | 6.44 | 41.7 | 2.81 | 4.01 |
| Plateau | 2013 | 619 | 11.0 | 14.2 | 2.17 | 2.44 |
| Plateau | 2018 | 437 | 19.2 | 18.5 | 2.34 | 2.33 |
| Plateau | 2024 | 876 | 8.56 | 33.2 | 2.23 | 2.34 |
| Rivers | 2013 | 498 | 20.7 | 40.4 | 2.83 | 3.83 |
| Rivers | 2018 | 378 | 18.0 | 40.7 | 3.03 | 3.99 |
| Rivers | 2024 | 720 | 10.3 | 20.3 | 3.03 | 4.12 |
| Sokoto | 2013 | 1198 | 8.51 | 1.34 | 1.17 | 2.06 |
| Sokoto | 2018 | 572 | 8.74 | 3.67 | 1.18 | 1.95 |
| Sokoto | 2024 | 1002 | 8.38 | 7.88 | 1.24 | 1.97 |
| Taraba | 2013 | 1123 | 18.2 | 7.66 | 1.71 | 2.18 |
| Taraba | 2018 | 592 | 14.7 | 8.78 | 1.98 | 2.16 |
| Taraba | 2024 | 636 | 10.4 | 9.75 | 1.70 | 2.12 |
| Yobe | 2013 | 972 | 8.96 | 1.34 | 1.27 | 1.81 |
| Yobe | 2018 | 652 | 13.2 | 1.84 | 1.32 | 1.63 |
| Yobe | 2024 | 837 | 31.3 | 8.24 | 1.38 | 1.92 |
| Zamfara | 2013 | 1193 | 7.21 | 3.52 | 1.18 | 1.71 |
| Zamfara | 2018 | 668 | 6.59 | 4.04 | 1.27 | 1.82 |
| Zamfara | 2024 | 677 | 7.24 | 19.4 | 1.30 | 2.34 |

**Source:** *Nigeria Demographic and Health Surveys (NDHS), 2013, 2018, 2024.*

Table 2 indicates that there is evidence of high spatiotemporal heterogeneity in the prevalence of pregnancy termination in Nigeria. To begin with, the increased rates were also focused on South-South (Rivers, 20.7%) and South-East (Imo, 18.3%), with the North-West (Kano, Katsina) being less prevalent. In the period between 2013 and 2018, there were both positive and negative trends, with such states as Abia and Bayelsa registering falls, whereas Jigawa and Gombe registered significant gains. The data assumes a nationwide wave as opposed to a local one by 2024. Other extreme prevalence rates of over 30 per cent were detected in Yobe, Bayelsa and Nasarawa and notable increases in Osun (28.8 per cent) and Kano (23.6 per cent). Conversely, Kwara and Niger were always outliers, with exceptionally low prevalence (1.5% and 6.2% respectively) despite huge samples. Although the same states (Kano, Katsina, Jigawa, and Bauchi) were the ones that reported the highest percentages of overall respondents, the high rates of termination were not necessarily related to the sample size. This failure of sample primacy points out important, state level contextual differences. The countrywide epidemic of 2018-2024 (both North and South) implies the radicalization of the reproductive situation. This pattern is probably a result of changing fertility choices, different contraceptive efficacies, more socioeconomic tension, or better reporting some time over the ten years.

**Table 2:** State-Level Distribution of Terminated pregnancy Counts and Rates by Year with proportion of total respondents (%) and Prevalence (%)

| Year | State | Total (N) | Yes | No | Proportion of Total Respondents (%) | Prevalence (%) |
| --- | --- | --- | --- | --- | --- | --- |
| 2013 | Abia | 469 | 42 | 427 | 1.7 | 9.0 |
| 2013 | Adamawa | 901 | 100 | 801 | 3.2 | 11.1 |
| 2013 | Akwa Ibom | 533 | 28 | 505 | 1.9 | 5.3 |
| 2013 | Anambra | 491 | 58 | 433 | 1.7 | 11.8 |
| 2013 | Bauchi | 1,259 | 226 | 1,033 | 4.5 | 18.0 |
| 2013 | Bayelsa | 763 | 84 | 679 | 2.7 | 11.0 |
| 2013 | Benue | 599 | 87 | 512 | 2.1 | 14.5 |
| 2013 | Borno | 572 | 40 | 532 | 2.0 | 7.0 |
| 2013 | Cross River | 490 | 61 | 429 | 1.7 | 12.4 |
| 2013 | Delta | 646 | 33 | 613 | 2.3 | 5.1 |
| 2013 | Ebonyi | 628 | 109 | 519 | 2.2 | 17.4 |
| 2013 | Edo | 568 | 48 | 520 | 2.0 | 8.5 |
| 2013 | Ekiti | 497 | 57 | 440 | 1.8 | 11.5 |
| 2013 | Enugu | 528 | 62 | 466 | 1.9 | 11.7 |
| 2013 | Gombe | 1,029 | 150 | 879 | 3.7 | 14.6 |
| 2013 | Imo | 437 | 80 | 357 | 1.6 | 18.3 |
| 2013 | Jigawa | 1,242 | 130 | 1,112 | 4.4 | 10.5 |
| 2013 | Kaduna | 826 | 85 | 741 | 2.9 | 10.3 |
| 2013 | Kano | 1,937 | 140 | 1,797 | 6.9 | 7.2 |
| 2013 | Katsina | 1,321 | 98 | 1,223 | 4.7 | 7.4 |
| 2013 | Kebbi | 1,043 | 73 | 970 | 3.7 | 7.0 |
| 2013 | Kogi | 462 | 20 | 442 | 1.6 | 4.3 |
| 2013 | Kwara | 637 | 25 | 612 | 2.3 | 3.9 |
| 2013 | Lagos | 885 | 158 | 727 | 3.1 | 17.9 |
| 2013 | Nasarawa | 597 | 58 | 539 | 2.1 | 9.7 |
| 2013 | Niger | 899 | 33 | 866 | 3.2 | 3.7 |
| 2013 | Ogun | 499 | 67 | 432 | 1.8 | 13.4 |
| 2013 | Ondo | 600 | 81 | 519 | 2.1 | 13.5 |
| 2013 | Osun | 537 | 20 | 517 | 1.9 | 3.7 |
| 2013 | Oyo | 625 | 73 | 552 | 2.2 | 11.7 |
| 2013 | Plateau | 619 | 68 | 551 | 2.2 | 11.0 |
| 2013 | Rivers | 498 | 103 | 395 | 1.8 | 20.7 |
| 2013 | Sokoto | 1,198 | 102 | 1,096 | 4.3 | 8.5 |
| 2013 | Taraba | 1,123 | 204 | 919 | 4.0 | 18.2 |
| 2013 | Yobe | 972 | 87 | 885 | 3.5 | 9.0 |
| 2013 | Zamfara | 1,193 | 86 | 1,107 | 4.2 | 7.2 |
| 2018 | Abia | 358 | 15 | 343 | 2.0 | 4.2 |
| 2018 | Adamawa | 548 | 45 | 503 | 3.1 | 8.2 |
| 2018 | Akwa Ibom | 307 | 42 | 265 | 1.7 | 13.7 |
| 2018 | Anambra | 504 | 28 | 476 | 2.8 | 5.6 |
| 2018 | Bauchi | 799 | 133 | 666 | 4.5 | 16.6 |
| 2018 | Bayelsa | 337 | 20 | 317 | 1.9 | 5.9 |
| 2018 | Benue | 523 | 41 | 482 | 2.9 | 7.8 |
| 2018 | Borno | 587 | 53 | 534 | 3.3 | 9.0 |
| 2018 | Cross River | 248 | 41 | 207 | 1.4 | 16.5 |
| 2018 | Delta | 292 | 18 | 274 | 1.6 | 6.2 |
| 2018 | Ebonyi | 565 | 57 | 508 | 3.2 | 10.1 |
| 2018 | Edo | 249 | 29 | 220 | 1.4 | 11.6 |
| 2018 | Ekiti | 279 | 36 | 243 | 1.6 | 12.9 |
| 2018 | Enugu | 332 | 23 | 309 | 1.9 | 6.9 |
| 2018 | Gombe | 701 | 139 | 562 | 3.9 | 19.8 |
| 2018 | Imo | 378 | 68 | 310 | 2.1 | 18.0 |
| 2018 | Jigawa | 754 | 147 | 607 | 4.2 | 19.5 |
| 2018 | Kaduna | 738 | 118 | 620 | 4.1 | 16.0 |
| 2018 | Kano | 1,054 | 137 | 917 | 5.9 | 13.0 |
| 2018 | Katsina | 832 | 132 | 700 | 4.7 | 15.9 |
| 2018 | Kebbi | 660 | 78 | 582 | 3.7 | 11.8 |
| 2018 | Kogi | 320 | 44 | 276 | 1.8 | 13.8 |
| 2018 | Kwara | 380 | 17 | 363 | 2.1 | 4.5 |
| 2018 | Lagos | 445 | 79 | 366 | 2.5 | 17.8 |
| 2018 | Nasarawa | 453 | 39 | 414 | 2.5 | 8.6 |
| 2018 | Niger | 666 | 103 | 563 | 3.7 | 15.5 |
| 2018 | Ogun | 293 | 19 | 274 | 1.6 | 6.5 |
| 2018 | Ondo | 299 | 13 | 286 | 1.7 | 4.3 |
| 2018 | Osun | 278 | 32 | 246 | 1.6 | 11.5 |
| 2018 | Oyo | 367 | 24 | 343 | 2.1 | 6.5 |
| 2018 | Plateau | 437 | 84 | 353 | 2.4 | 19.2 |
| 2018 | Rivers | 378 | 68 | 310 | 2.1 | 18.0 |
| 2018 | Sokoto | 572 | 50 | 522 | 3.2 | 8.7 |
| 2018 | Taraba | 592 | 87 | 505 | 3.3 | 14.7 |
| 2018 | Yobe | 652 | 86 | 566 | 3.7 | 13.2 |
| 2018 | Zamfara | 668 | 44 | 624 | 3.7 | 6.6 |
| 2024 | Abia | 470 | 73 | 397 | 1.8 | 15.5 |
| 2024 | Adamawa | 789 | 204 | 585 | 3.0 | 25.9 |
| 2024 | Akwa Ibom | 494 | 140 | 354 | 1.9 | 28.3 |
| 2024 | Anambra | 643 | 169 | 474 | 2.5 | 26.3 |
| 2024 | Bauchi | 884 | 108 | 776 | 3.4 | 12.2 |
| 2024 | Bayelsa | 513 | 160 | 353 | 2.0 | 31.2 |
| 2024 | Benue | 631 | 118 | 513 | 2.4 | 18.7 |
| 2024 | Borno | 671 | 72 | 599 | 2.6 | 10.7 |
| 2024 | Cross River | 549 | 117 | 432 | 2.1 | 21.3 |
| 2024 | Delta | 613 | 98 | 515 | 2.4 | 16.0 |
| 2024 | Ebonyi | 840 | 157 | 683 | 3.2 | 18.7 |
| 2024 | Edo | 541 | 48 | 493 | 2.1 | 8.9 |
| 2024 | Ekiti | 369 | 96 | 273 | 1.4 | 26.0 |
| 2024 | Enugu | 485 | 69 | 416 | 1.9 | 14.2 |
| 2024 | Gombe | 806 | 150 | 656 | 3.1 | 18.6 |
| 2024 | Imo | 653 | 178 | 475 | 2.5 | 27.3 |
| 2024 | Jigawa | 955 | 118 | 837 | 3.7 | 12.4 |
| 2024 | Kaduna | 1,080 | 240 | 840 | 4.2 | 22.2 |
| 2024 | Kano | 1,210 | 285 | 925 | 4.7 | 23.6 |
| 2024 | Katsina | 914 | 111 | 803 | 3.5 | 12.1 |
| 2024 | Kebbi | 1,008 | 199 | 809 | 3.9 | 19.7 |
| 2024 | Kogi | 699 | 58 | 641 | 2.7 | 8.3 |
| 2024 | Kwara | 664 | 10 | 654 | 2.6 | 1.5 |
| 2024 | Lagos | 631 | 115 | 516 | 2.4 | 18.2 |
| 2024 | Nasarawa | 750 | 234 | 516 | 2.9 | 31.2 |
| 2024 | Niger | 935 | 58 | 877 | 3.6 | 6.2 |
| 2024 | Ogun | 670 | 148 | 522 | 2.6 | 22.1 |
| 2024 | Ondo | 483 | 56 | 427 | 1.9 | 11.6 |
| 2024 | Osun | 434 | 125 | 309 | 1.7 | 28.8 |
| 2024 | Oyo | 745 | 48 | 697 | 2.9 | 6.4 |
| 2024 | Plateau | 876 | 75 | 801 | 3.4 | 8.6 |
| 2024 | Rivers | 720 | 74 | 646 | 2.8 | 10.3 |
| 2024 | Sokoto | 1,002 | 84 | 918 | 3.9 | 8.4 |
| 2024 | Taraba | 636 | 66 | 570 | 2.5 | 10.4 |
| 2024 | Yobe | 837 | 262 | 575 | 3.2 | 31.3 |
| 2024 | Zamfara | 677 | 49 | 628 | 2.6 | 7.2 |

**Source:** *Nigeria Demographic and Health Surveys (NDHS), 2013, 2018, 2024.*

Figure 1 indicates that there is an evident temporal increase in the rates of terminated pregnancies and spatial polarization among the Nigerian states during the years 2013 to 2024. Generally, in 2013, there was low to moderate rates with minimal spatial clustering, especially in the majority of northern states (e.g., Sokoto, Zamfara, Katsina, Jigawa, Yobe, Borno, Bauchi) and was indicative of high socio-cultural constraint, low access to services, and underreporting. The moderate levels were clumped in sections of the South-West, South-South and North-Central region, which showed the initial fertility transition. By 2018, spatial differentiation was intensified, where a developing high-rate belt was observed in the southern and urbanized states, including Lagos, Rivers, Delta, Bayelsa, Akwa Ibom, Ogun, Osun, Oyo and Cross River, as well as increase in the rates in the South-East and some parts of the North-Central zone. By contrast, the majority of North-West and North-East states were low but some new rises in Kano, Kaduna, Adamawa and Gombe were indicative of increased reproductive stress. By 2024, the trend had developed into very polarized clusters, whereby adjacent high-intensity areas were located in the South-West, South-South, South-East, and some areas of the North-Central zone, which is in line with the increased fertility transition, the increased experimentation with contraceptives, and the delayed marriage and better reporting. Most northern states simply stayed down, but with rising gradual gains, especially in Kano, Kaduna, Niger, Adamawa and Gombe, are probably indicative of unmet contraceptive demand and structural constraints as opposed to reproductive alternative.

**Figure 1:**
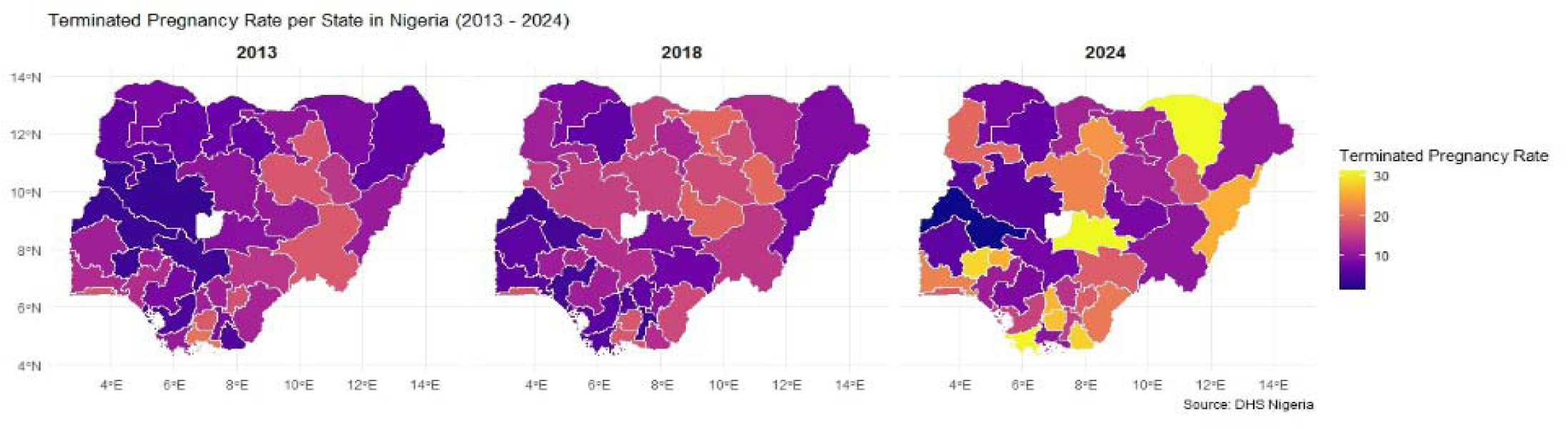
Shifting Geography of Terminated Pregnancy Rates in Nigeria: 2013, 2018 and 2024 DHS Evidence

**Figure 2:**
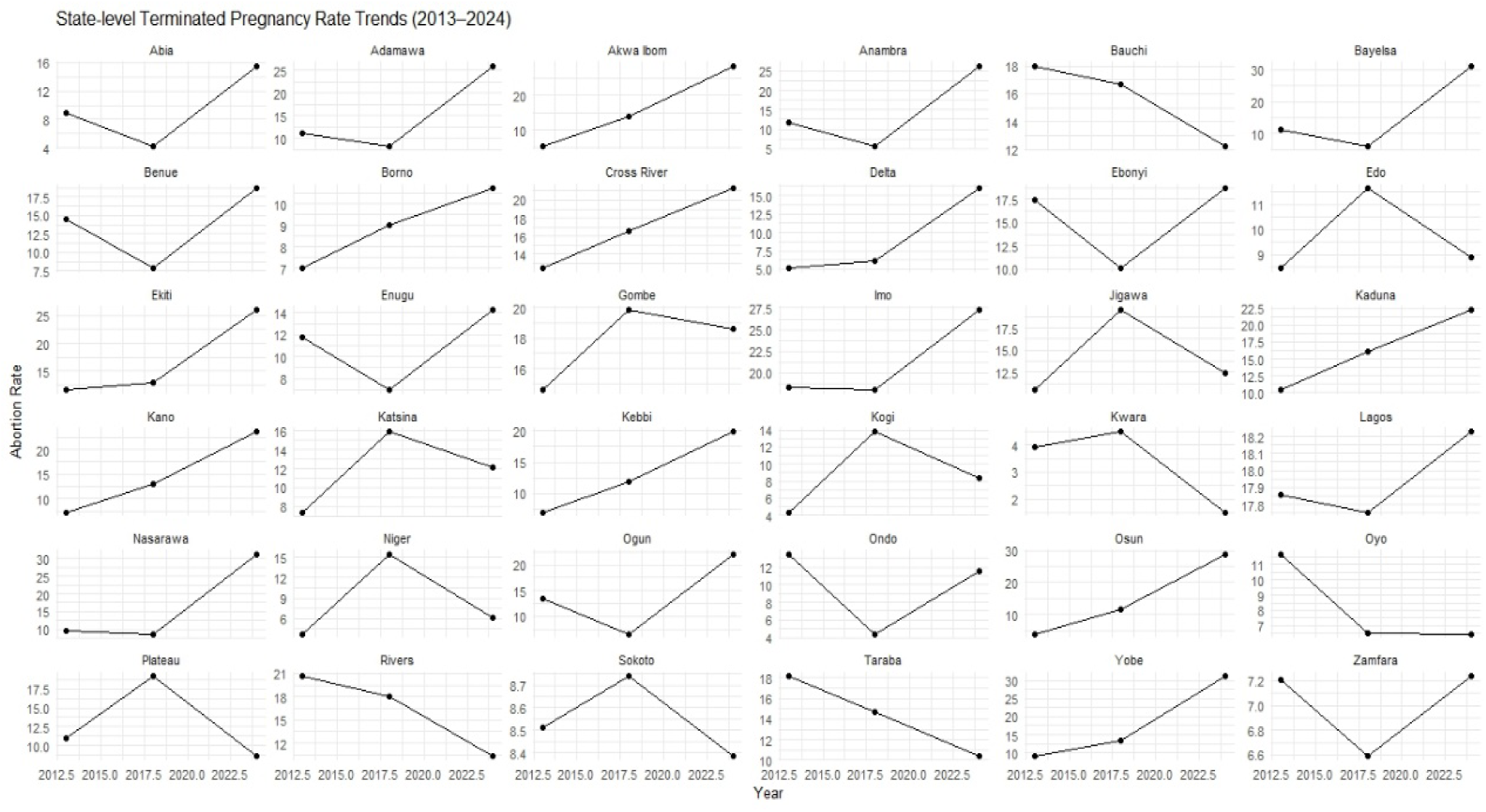
Divergent State-Level Trends in Terminated Pregnancy Rates in Nigeria (2013–2024)

The spatiotemporal dynamic of pregnancy termination in Nigeria suggests a shift of a steady, locally determined plateau in the year 2013 to a highly polarized country of 2024, with two diverse regional drivers. Acute upward curves in the Southern and Coastal states, such as Lagos, Bayelsa, Akwa Ibom and Osun, arose as a proactive fertility-management approach, termination, a supplement to modern contraception among the educated women in the Southern and Coastal states as they negotiated delayed childbearing and an expanded reproductive freedom. In contrast, the Northern and North-Eastern region (headed by Yobe, Adamawa, and Kano) were under the influence of crisis-related surges due to the high unmet contraceptive need, socioeconomic instability, and structural impediments but not reproductive choice. The South-East was high volatility due to economic pressures and the North-Central was a buffer area, though Kwara was still an outlier of its own as it fell to a national low of 1.5%. Finally, as this disparity between the choice-oriented South and the vulnerability-oriented North keeps expanding, it confirms that pregnancy termination ceased to be a single consequence of disadvantage and requires state-based policies addressing reproductive health challenges, as opposed to a nationally uniform solution.

According to BYM2 model, incidence of Terminated pregnancy in Nigeria by the state is highly heterogeneous, both in time, and socioeconomic. It is indicated in table 3.4 that increased average state wealth was significantly and credibly connected to the number of Terminated pregnancies indicating that there was a protecting socioeconomic effect, which was present even after the account was made against spatial dependence. The mean education showed a negative albeit very inaccurate association where literacy showed a significant effect with wide credible range of values that showed higher reporting or access of services in more literate states than were in a causal association. The rate of contraceptive use showed a poor positive relationship with the number of Terminated pregnancies that showed access to reproductive health service and reporting instead of a greater risk.

**Table 3.4:** Fixed Effects Estimates from the BYM2 Model of State-Level Terminated pregnancy Counts (2013)

| Covariate | Mean | SD | 2.5%<br>Quantile | Median | 97.5%<br>Quantile | Mode | KLD($\times 10^{-3}$) |
| --- | --- | --- | --- | --- | --- | --- | --- |
| (Intercept) | 0.1195 | 0.4944 | −0.8296 | 0.1099 | 1.1234 | 0.1101 | 6.95 |
| avg_education | −0.5419 | 0.9817 | −2.4828 | −0.5402 | 1.3892 | −0.5402 | 3.07 |
| avg_wealth | −0.8930 | 0.2444 | −1.3823 | −0.8902 | −0.4189 | −0.8902 | 4.27 |
| contraceptive_use_rate | 0.0161 | 0.0106 | −0.0048 | 0.0161 | 0.0370 | 0.0161 | 2.91 |
| avg_literacy | 1.6248 | 1.1480 | −0.6327 | 1.6227 | 3.8945 | 1.6227 | 3.02 |

**Source:** Nigeria Demographic and Health Surveys (NDHS), 2013, 2018, 2024.

**Table 3.5:** Fixed Effects Estimates from the BYM2 Model of State-Level Terminated pregnancy Counts (2018)

| Covariate | Mean | SD | 2.5%<br>Quantile | Median | 97.5%<br>Quantile | Mode | KLD($\times 10^{-3}$) |
| --- | --- | --- | --- | --- | --- | --- | --- |
| (Intercept) | 0.4515 | 0.5623 | −0.7123 | 0.4679 | 1.5204 | 0.4668 | 11.05 |
| avg_education | 0.2041 | 0.4402 | −0.6582 | 0.2019 | 1.0784 | 0.2020 | 3.23 |
| avg_wealth | −0.3459 | 0.2644 | −0.8691 | −0.3453 | 0.1736 | −0.3453 | 3.02 |
| contraceptive_use_rate | 0.0161 | 0.0092 | −0.0020 | 0.0161 | 0.0341 | 0.0161 | 2.93 |
| avg_literacy | −0.2155 | 0.8273 | −1.8445 | −0.2162 | 1.4171 | −0.2161 | 2.91 |

**Source:** Nigeria Demographic and Health Surveys (NDHS), 2013, 2018, 2024.

**Table 3.6:** Fixed Effects Estimates from the BYM2 Model of State-Level Terminated pregnancy Counts (2024)

| Variable | Mean | SD | 2.5%<br>Quantile | Median<br>(50%) | 97.5%<br>Quantile | Mode | KLD($\times 10^{-3}$) |
| --- | --- | --- | --- | --- | --- | --- | --- |
| Intercept | −0.8101 | 0.7051 | −2.2796 | −0.7864 | 0.5192 | −0.7884 | 11.46 |
| Average education | 0.0144 | 0.6691 | −1.3010 | 0.0122 | 1.3429 | 0.0125 | 3.14 |
| Average wealth | −0.3791 | 0.3158 | −0.9945 | −0.3815 | 0.2498 | −0.3817 | 3.62 |
| Contraceptive use rate | 0.0145 | 0.0110 | −0.0070 | 0.0145 | 0.0362 | 0.0145 | 2.99 |
| Average literacy | 0.9107 | 1.0013 | −1.0655 | 0.9114 | 2.8828 | 0.9114 | 3.08 |

**Source:** Nigeria Demographic and Health Surveys (NDHS), 2013, 2018, 2024.

Table 3.5 indicates changing socioeconomic links with time. By 2018, the protective association with wealth became smaller and credible intervals were found to cross zero, indicating that socioeconomic convergence occurred in terminated pregnancy. Education moved with a positive correlation meaning that it was more incidence in the more educated states. A less significant negative correlation was observed with literacy, which may be evidence of greater reproductive autonomy. The use of contraceptives also had a positive correlation with time, which is in tandem with its propensity to be a marker of healthcare access as opposed to a direct risk factor.

Table 3.6 reveals that there was a reduction in the baseline risk of terminated pregnancy by state with wealth providing a weak, re-emerging protective effect. Neither education nor literacy had a significant effect independently; literacy once again had a positive effect-indicating that its effect may be at times situational or age mediated. The use of contraceptives had a small yet significant positive correlation over the time. In general, the most predictable structural variable was wealth, but education and literacy showed different patterns throughout the analysis.

As can be seen in figure 4, the BYM2 model indicates the evident temporal and socioeconomic heterogeneity in the occurrence of pregnancy termination in Nigerian states. The 2013 report (averaging state wealth) has delivered a substantial and effective protective impact. Nevertheless, this protective power has fallen by 2018 as there is no longer a wide incidence gap between the socioeconomic layers with a small negative wealth effect reappearing in 2024. This change was reflected in educational attainment, with an imprecise negative relationship in 2013 and positive association in 2018 that indicates that higher education is becoming increasingly linked to reproductive choices and termination. Over the decade, literacy and contraceptive use had positive relationships with the number of terminations; these relations do not point at increased risk, but the associations have been in the positive direction that implies better access to services and more appropriate reporting in states with a higher quality of reproductive health infrastructure. Finally, these changing determinants have formed a two-polar reproductive regime where wealth is still a fixed structural component, but education and literacy have been transformed into the main agents of active reproductive control as it is shown on the spatiotemporal maps in figure above.

**Table 3.7:** Fixed Effects (IRRs) for BYM2 Models, 2013 - 2024.

| Model | Year | Predictor | Mean (log) | IRR | 95% CI (Lower) | 95% CI (Upper) |
| --- | --- | --- | --- | --- | --- | --- |
| BYM2 | 2013 | Intercept | 0.120 | 1.127 | 0.436 | 3.075 |
| BYM2 | 2013 | Avg. education | −0.542 | 0.582 | 0.084 | 4.012 |
| BYM2 | 2013 | Avg. wealth | −0.893 | 0.409 | 0.251 | 0.658 |
| BYM2 | 2013 | Contraceptive use rate | 0.016 | 1.016 | 0.995 | 1.038 |
| BYM2 | 2013 | Avg. literacy | 1.625 | 5.078 | 0.531 | 49.128 |
| BYM2 | 2018 | Intercept | 0.451 | 1.571 | 0.490 | 4.570 |
| BYM2 | 2018 | Avg. education | 0.204 | 1.226 | 0.518 | 2.940 |
| BYM2 | 2018 | Avg. wealth | −0.346 | 0.708 | 0.419 | 1.190 |
| BYM2 | 2018 | Contraceptive use rate | 0.016 | 1.016 | 0.998 | 1.030 |
| BYM2 | 2018 | Avg. literacy | −0.216 | 0.806 | 0.158 | 4.120 |
| BYM2 | 2024 | Intercept | −0.810 | 0.445 | 0.102 | 1.680 |
| BYM2 | 2024 | Avg. education | 0.014 | 1.014 | 0.272 | 3.830 |
| BYM2 | 2024 | Avg. wealth | −0.379 | 0.685 | 0.370 | 1.280 |
| BYM2 | 2024 | Contraceptive use rate | 0.014 | 1.015 | 0.993 | 1.040 |
| BYM2 | 2024 | Avg. literacy | 0.911 | 2.486 | 0.345 | 17.860 |

**Source:** *Nigeria Demographic and Health Surveys (NDHS), 2013, 2018, 2024.*

**Table 3.8:** Model Diagnostics BYM2 Models, 2013 - 2024.

| Model | DIC | WAIC |
| --- | --- | --- |
| BYM2 (2013) | 289.86 | 284.11 |
| BYM2 (2018) | 275.73 | 270.27 |
| BYM2 (2024) | 305.42 | 297.97 |

**Source:** *Nigeria Demographic and Health Surveys (NDHS), 2013, 2018, 2024.*

**Figure 4:**
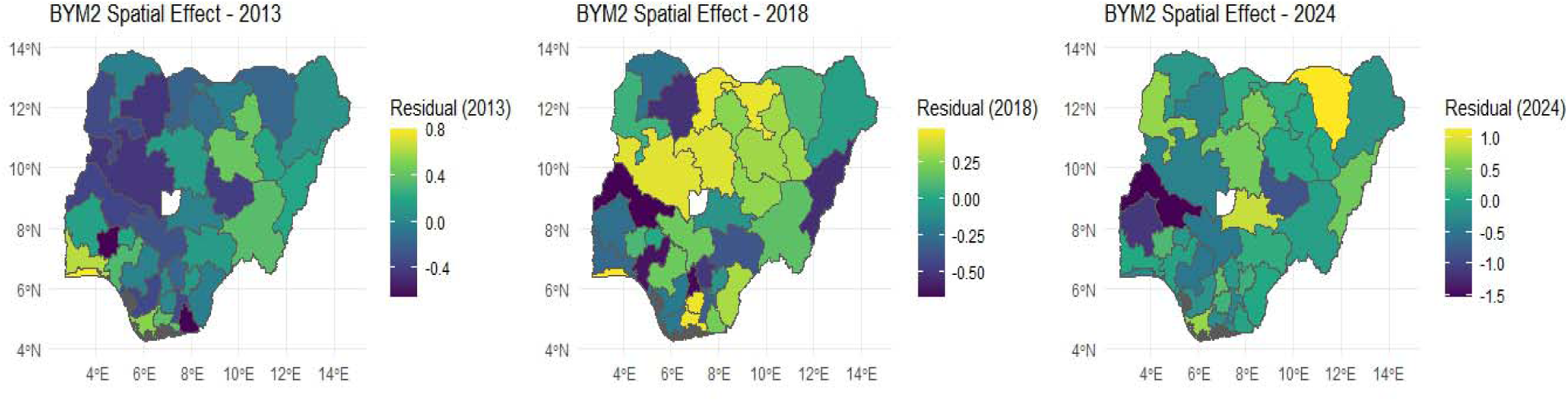
State-Level Socio-Demographic Determinants of Abortion Counts: BYM2 Model Estimates for 2013 and 2018

Model Diagnostics and Fixed Effects (IRRs) for BYM2 Models, 2013 - 2024

According to Table 3.7 of the 2013, 2018, and 2024 BYM2 state-level counts of abortion, the common fixed effects are that average wealth is negatively related to counts of Terminated pregnancy (IRRs < 1) are, and the rates of contraceptive use have a smaller but still positive relationship (IRRs slightly > 1) with reported counts of Terminated pregnancy. The effect of average education and literacy is more time-varying: in 2013, higher literacy has a very strong positive effect on the number of abortions (IRR = 5.08), in 2018 it is negative (IRR = 0.81), and in 2024 again it is positive (IRR = 2.49), and the effect of these socio-demographic variables changes over time. Intercepts confirm the fact that the number of Terminated pregnancies at baseline varies by year with the highest numbers in 2018 (IRR = 1.57) and the lowest numbers in the year 2024 (IRR = 0.45). As it is stated in table 3.8, it is possible to mention that the model diagnostics proves the good fit of BYM2 as the values of DIC and WAIC in 2013 were (289.86 /284.11), in 2018, (305.42/ 297.97), and the models could be used as a good predictor of the spatial heterogeneities in the state level and also describe the relative risks of the major covariates.

## 4.0 CONCLUSION

This paper presents a full spatio-temporal evaluation of the patterns of terminated pregnancies in Nigeria, showing that there were significant regional disparities and explicit structural change between 2013 and 2024. According to the findings, the trends in Nigeria have exemplified a transition between the comparatively diffuse regional variation to a highly polarized reproductive health environment. Southern states are becoming more indicative of an advanced fertility transition, in which terminated pregnancy becomes a self-professed and complementary fertility-regulation tactic in combination with increasing education, wealth, and access to contraceptives. Northern Nigeria, by contrast, is still typified by reproductive vulnerability, with rises in terminated pregnancy being more effectively linked to unmet contraceptive need, socioeconomic turbulence, early matrimony, and structural disadvantages to reproductive healthcare in comparison to autonomous reproductive choice. The BYM2 modeling framework using Bayesian hierarchical spatio-temporal modeling system proves that these discrepancies are not accidental but are influenced by existing spatial dependence and changing socioeconomic gradients. Although the first effect of household wealth is the protective one, over time, education and literacy have become the most potent factors on reproductive decisions and reporting, which underpins the contribution of human capital to reproductive outcomes. These findings not only dispute mono-fits-all national strategies but also demonstrate the need to have decentralized and state-specific reproductive health policies that would effectively and sustainably respond to autonomy-based termination in the South and vulnerability-based termination in the North. Policies should consequently strive to expand structural contraceptive and maternal health provision in the North and South should capitalize on education-based interventions to maximize contraceptive use, and minimize unwanted fertility. To strengthen further the evidence-based planning, in future studies, it is recommended that migration and displacement dynamic be added to spatio-temporal models since population mobility is increasingly redefining reproductive risk among fast urbanizing southern states and vulnerable northern areas.

## Statements and Declarations

### Author Contributions

**Taiwo Oyewole Asifat** conceptualized and designed the study, led the literature synthesis, and performed the initial drafting of the manuscript. He was responsible for defining the socio-ecological framework used to analyze reproductive health disparities and managed the overall integration of the epidemiological discussion. **Oluwafemi Lawal Bisiriyu** was responsible for data curation, developed the Bayesian spatio-temporal methodology, performed the formal statistical analysis, generated the visualizations, and served as the corresponding author overseeing the submission process. **Ogunetimoju Abolaji Moses** provided critical interpretation of the results and conducted a rigorous final review and editing of the manuscript to ensure methodological consistency and intellectual quality. All authors have read and approved the final manuscript for publication.

## Clinical trial number

Not applicable.

## Institutional Review Board Statement

This methodological study utilized de-identified, publicly available secondary datasets. As such, no additional ethical approval was required beyond the original data collection protocols.

## Data Availability

All data produced in the present study are available upon reasonable request to the authors
All data produced in the present work are contained in the manuscript
All data produced are available online at https://dhsprogram.com/data/available-datasets.cfm

## Acknowledgments

The authors would like to express their gratitude to The DHS Program (ICF International, Rockville, MD, USA) for granting access to the 2013, 2018, and 2024 Nigeria Demographic and Health Survey (NDHS) datasets used in this study. We also acknowledge the National Population Commission (NPC) of Nigeria for their role in the successful implementation of the surveys and for ensuring the availability of high-quality, nationally representative data. Special thanks are extended to the fieldworkers and the thousands of women across Nigeria who participated in these surveys, providing the essential data that makes this reproductive health research possible. Finally, we thank the reviewers for their insightful comments that helped refine the Bayesian spatio-temporal framework presented in this work.

